# Association of an Aquaporin-4 Haplotype with Cognition, Brain Volume, and Dementia Risk in Community-Dwelling Individuals without Dementia

**DOI:** 10.64898/2025.12.15.25342248

**Authors:** Emma L. Palatsides, Dibya Himali, Lachlan Cribb, Gina M. Peloso, Joanne Ryan, Qiong Yang, Charles S. DeCarli, Alexa S. Beiser, Sudha Seshadri, Marina G. Cavuoto, Stephanie Yiallourou, Andrée-Ann Baril, Jayandra J. Himali, Matthew P. Pase

**Affiliations:** Turner Institute for Brain and Mental Health, School of Psychological Sciences, Monash University, Melbourne, VIC, Australia; Framingham Heart Study, Framingham, MA, USA; Department of Neurology, Boston University Chobanian & Avedisian School of Medicine, Boston, MA, USA; Department of Biostatistics, Boston University School of Public Health, Boston, MA, USA; School of Public Health and Preventive Medicine, Monash University, Melbourne, VIC, Australia; Department of Neurology & Imaging of Dementia and Aging Laboratory, University of California, Davis, Davis, CA, USA; Glenn Biggs Institute for Alzheimer’s & Neurodegenerative Diseases, San Antonio, TX, USA; University of Texas Health San Antonio, San Antonio, TX, USA; National Ageing Research Institute, Melbourne, VIC, Australia; Center for Advanced Research in Sleep Medicine, Hôpital du Sacré-Coeur de Montréal, CIUSSS-NIM, Montreal, QC, Canada; Department of Medicine, Université de Montréal, Montreal, QC, Canada

## Abstract

**Background and Objectives:** The aquaporin-4 (AQP4) water channel plays an integral role in clearing brain waste. However, little is known about whether variations in the *AQP4* gene contributes to brain health or dementia risk. We aimed to determine whether a functional *AQP4* haplotype was associated with cognition, brain volumes, or incident dementia.

**Methods:** This study included participants from two prospective cohort studies. Firstly, participants from the Original, Offspring, New Offspring Spouse, and Generation 3 cohorts from the Framingham Heart Study (FHS; enrolled 1948-2005) were included with dementia follow-up until 2022. Original FHS participants were from Framingham, Massachusetts at the time of enrollment. Analyses were replicated for brain MRI outcomes and incident dementia with UK Biobank participants (enrolled 2006-2010) who were followed until 2022. All participants were dementia-free at the time of cognitive assessment, brain MRI, and commencement of dementia follow-up. Linear regression models were conducted to analyze the associations between *AQP4* and the cognitive and brain MRI outcomes. Cox proportional hazard regression models were conducted to analyze the association between *AQP4* and incident all-cause dementia risk. Data analysis spanned February 2023 to July 2025.

**Results:** The FHS sample comprised 3,847 participants (65% homozygote major, 31% heterozygote, 4% homozygote minor) with cognitive testing (mean age 61±11 years; 54% women); 3,332 had brain MRI. Heterozygotes displayed better verbal episodic memory (β[95% CI], 0.32 [0.09, 0.55], *p*=0.007, N=1,201) and larger hippocampal volumes (β[95% CI], 0.09 [0.02, 0.16], *p*=0.012, N=1,052) compared to homozygote majors. Similar findings were observed in the UK Biobank; homozygote minors displayed larger hippocampal volumes (β[95% CI], 0.05 [<0.01, 0.11], *p*=0.035, N=32,219) and lower amounts of DTI measured free water (β[95% CI], -0.09 [-0.16, - 0.03], *p*=0.005, N=31,807) compared to homozygote majors. Heterozygotes displayed a statistically significant lower rate of incident all-cause dementia (HR=0.93, 95% CI [0.88, 0.98], *p*=0.012, N=114,868, incident cases=5,625) compared to homozygote majors.

**Discussion:** Carrying at least one minor allele at an *AQP4* haplotype (homozygote minors or heterozygotes) was linked to better verbal episodic memory, larger hippocampal volumes, lower amounts of free water, and lower dementia risk. Further studies are required to replicate these results amongst diverse samples.

## INTRODUCTION

Aquaporin-4 (AQP4) is the predominant transmembrane protein in the central nervous system that permits the diffusion of water and small particles across cell membranes.^1^ AQP4 plays an essential role in the glymphatic system by facilitating fluid influx into the brain parenchyma from the periarterial space, which subsequently flows in a directionally polarized fashion, carrying waste products into the perivenous space prior to being cleared from the brain.^2^ Additionally, its role in water regulation has important implications for how the brain responds to neuroinflammation and insult. For example, AQP4 channels can be up or down regulated in conditions that involve a neuroinflammatory response, such as neuromyelitis optica, stroke, and brain injury.^3–5^

Neuroinflammation and a breakdown of clearance mechanisms may increase the risk of Alzheimer’s disease dementia,^6,7^ which involves the gradual accumulation of fibrillated β-amyloid (Aβ) and hyperphosphorylated tau proteins in the brain.^8^ For AQP4 specifically, deletion of its encoding gene has been associated with substantial increases in Aβ and tau in mice.^9^ In humans, loss of AQP4 perivascular localization (AQP4 proteins that have not anchored to the correct location on an astrocyte) has been associated with greater Aβ burden in individuals with AD.^10^ More recently, combinations of *AQP4* single nucleotide polymorphisms (SNPs) have been linked to higher Aβ burden across two cohorts.^11^ Alterations in the functioning of AQP4 channels present at the blood-brain barrier (BBB) may also impact glymphatic function, neuronal health, and brain homeostasis.^12–14^

Despite these mechanisms, few studies have explored the relationship between *AQP4* genetic variation and dementia or its endophenotypes.^15,16^ One study in young adults found carrying the low AQP4-expressing haplotype (homozygote for the minor allele or heterozygote) was associated with better reaction times following sleep deprivation compared to homozygote majors.^17^ Carrying the minor allele at this haplotype has a population prevalence of approximately 20% and is associated with a reduced expression of AQP4 channels compared to homozygote majors.^17,18^ Although this seems counterintuitive, it is thought that the minor allele may augment slow wave sleep—and therefore cognitive performance—to compensate for a reduction in AQP4 channel expression.^17^

We aimed to explore the relationship between an *AQP4* haplotype with cognition, brain volumes, and incident all-cause dementia risk. Since carrying the minor allele at the *AQP4* haplotype has previously been associated with better cognition,^17^ it was hypothesized that carrying at least one minor allele, compared to carrying two major alleles, would result in better cognitive performance, larger brain volumes, and a lower rate of dementia.

## METHODS

### Participants

Participants were from the Framingham Heart Study (FHS), a multigenerational prospective community-based cohort study that commenced in 1948. The Original cohort comprised 5,209 participants, who were residents of Framingham, Massachusetts at the time of enrollment.^19^ The Offspring cohort was established in 1971 and included 5,124 participants who were either the biological children, adoptive children, or spouses of the Original cohort.^20^ The Generation 3 cohort commenced in 2002 and included 4,095 children of the Offspring cohort.^21^ An additional cohort, named New Offspring Spouse, commenced in 2003 and included 103 spouses of the Offspring cohort.^21^ Surviving participants continue to be followed at regular examination cycles and are under continuous surveillance for medical events, such as myocardial infarction, stroke, and dementia.^19^

For the present study, we included Original, Offspring, New Offspring Spouse, and Generation 3 cohorts for the cognition and MRI analyses. We used MRI and cognitive assessment results that were proximal to exam 26 for the Original cohort (1999–2002), exam 7 for the Offspring cohort (1999-2005), and exam 2 for New

Offspring Spouse (2010–2015) and Generation 3 (2009–2014) cohorts. All covariates were obtained from those exam cycles. Only Original and Offspring cohort participants were included in the dementia risk analyses because Generation 3 participants are substantially younger and New Offspring Spouse have shorter follow-up periods. Only Offspring, New Offspring Spouse, and Generation 3 cohorts had Diffusion Tensor Imaging (DTI) completed. For the outcomes of cognition and brain MRI, we excluded participants who were younger than 45 years at the time of their outcome assessment. For incident dementia risk, participants were aged 65 ± 2.5 years at the commencement of dementia follow-up. Participants with dementia or unknown dementia status were excluded at study entry (for dementia risk analyses) or the time of their outcome assessment (for cognition and brain MRI analyses). Sample selection information for each analysis is presented in eFigures 1-3.

**Figure 1.**
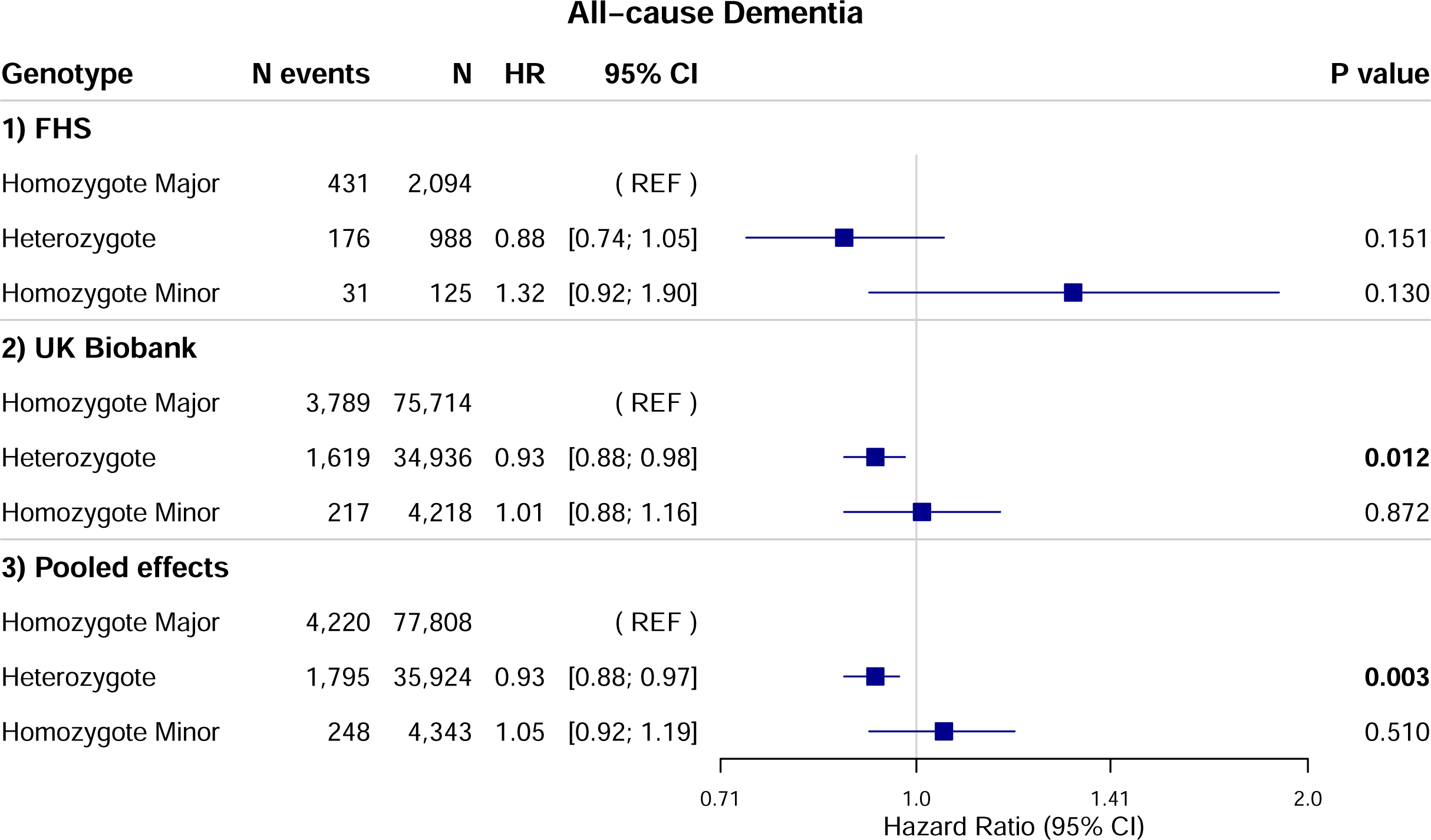
AQP4 and Dementia Risk Forest Plot. *Note. FHS:* Average follow-up time was 20 years (SD=7). Results were adjusted for entry age, sex, cohort (Original and Offspring), and *APOE* (ε4 carrier versus non-carrier). *UK Biobank:* Average follow-up time was 13 years (SD=3). Results were adjusted for age, sex, ethnicity (white, other), and *APOE* (ε4 carrier versus non-carrier). *Pooled Effects*: Pooled effects was calculated in a meta-analysis using the inverse variance method. Bold indicates statistical significance, *p*<.05; HR=hazard ratio; CI=confidence interval; FHS=Framingham Heart Study

### Genotyping of AQP4

Genotyping of participant DNA was completed at the Affymetrix Research Services Laboratory (ARSL) in Santa Clara, California, USA. Peripheral blood samples were stored at -80°C and genotyping was completed using Affymetrix GeneChip Human Mapping 500K Array and the 50K Human Gene Focused Panel. Samples were excluded if a participant call rate was less than 97%, a per-subject heterozygosity was ±5 SD from the mean, or a per-subject number of Mendelian errors was greater than 165 (99th quantile). Genotyping from 433,510 SNPs in 8,481 individuals passed these quality-control measures. The BRLMM (Bayesian Robust Linear Model with Mahalanobis distance) algorithm was used for allele calling. Principal-components analysis was applied to evaluate population structure (infer axes of variation) using a subset of 425,173 SNPs with minor allele frequency (MAF)≥0.01, Hardy-Weinberg equilibrium (HWE) p≥10^−6^, and call rate≥0.95.

Following previously reported methods,^17^ we examined genetic variation of *AQP4* by calculating an *AQP4* haplotype. Studying a haplotype allowed us to examine a combination of alleles inherited together rather than considering them in isolation.

The haplotype consists of 8 SNPs: rs162007, rs162008, rs63514, rs455671, rs335931, rs335930, rs335929, and rs16942851. Of these, three were included as tag SNPs to represent the haplotype (rs335931, rs335929, and rs16942851), as they have high linkage disequilibrium.^17^ We classified participants into one of the following three groups based on the number of *AQP4* minor alleles present: homozygote major (no minor alleles for all three SNPs); heterozygote (one minor allele at each SNP); or homozygote minor (two minor alleles at each SNP; see Table 1).

**Table 1.** Classification of haplotype based on *AQP4* tag SNPs

| SNP | Homozygote Major | Heterozygote | Homozygote Minor |
| --- | --- | --- | --- |
| rs335931 | AA | AG | GG |
| rs335929 | AA | AC | CC |
| rs16942851 | TT | TG | GG |

*Note.* Participants were classified into one of the following haplotype groups based on the number of minor alleles present at each SNP: homozygote major (no minor alleles for all three SNPs); heterozygote (one minor allele at each SNP); or homozygote minor (two minor alleles at each SNP). The alleles for each SNP are presented above (e.g., the major allele is A and the minor allele is G for rs335931).

Approximately 99.84% of FHS participants were classified into one of these three groups. Six participants were excluded from the cognition and dementia risk samples and five were excluded from the MRI sample for having a rare *AQP4* haplotype (e.g. a combination of major/minor homozygotes for one allele but not the others).

Previous research has identified that homozygote minors and heterozygotes have a lower expression of AQP4 channels compared to homozygote majors.^18^ Consequently, studies have tended to classify participants into two groups: minor allele carrier group (low AQP4 expression; homozygote minor or heterozygote) or homozygote major (high AQP4 expression).^17^ This is also to additionally overcome the low percentage of participants who belonged to the homozygote minor genotype group. However, given our large sample size, we explored whether there were any differences between patterns of relationships or effect sizes when carrying one *AQP4* minor allele versus two.

### Cognitive Assessment

Participants underwent comprehensive neuropsychological testing administered by trained research assistants. The following cognitive tests were included: Logical Memory immediate and delayed recall for verbal episodic memory, Visual Reproduction immediate and delayed recall for visual memory, Trail Making Test B minus A for cognitive flexibility, and Similarities for abstract reasoning.

### Brain Volumes

The MRI sample was a subset of the cognition sample (87% of participants with cognition also had MRI; eFigures 1 and 2). The FHS MRI protocol has been described previously.^22,23^ Briefly, participants were imaged across several MRI scanners with field strength ranging from 1 to 3T. A 3-dimensional T1-weighted and 2 or 3-dimensional FLAIR imaging sequences were used. All images were transferred to the University of California Davis Medical Center for processing, where staff were blinded to clinical information. Automated procedures with quality control were used for the segmentation and quantification of brain volumes. MRI image analyses were completed by trained operators under supervision of CD. A convolutional neural network method was used to approximate total cerebral cranial volume.^24^ Images were further segmented into 4 tissue types (gray matter, white matter, CSF, and white matter hyperintensity volumes) using previously published methods.^25–27^

Hippocampal segmentation was conducted using an atlas-based diffeomorphic method with a minor label-refinement modification.^28,29^ Non-linear co-registration to the DKT atlas was used to derive regional grey matter volumes.^29–31^ Free water maps were coregistered.^32^ Each voxel corresponded to the same brain region for each participant.^32^

The following brain volumes were included for analysis: hippocampal volume, white matter hyperintensities volume, cortical grey matter volume, and total brain volume. All volumes were corrected for differences in head size by computing each variable as a percentage of total intracranial volume. Free water was assessed using DTI to capture subtle differences in white matter integrity and fluid flow.

### Dementia Case Ascertainment

Participants in the FHS are under continuous surveillance for dementia. The dementia status participants is determined by a panel comprised of at least one neurologist and one neuropsychologist. Cognitive screening of Original and Offspring participants is conducted using the Mini-Mental State Examination (MMSE) at each examination cycle. Participants are flagged as having possible cognitive impairment if their MMSE score falls below education-based cut-off scores, if there has been a decline of 3 or more points between examinations, or if there has been a decrease of 5 or more points from the participant’s highest MMSE score documented thus far.

Additionally, participants can be flagged with suspected cognitive impairment if concerns are raised following discussions with the participant or their relatives, or from evidence gathered from hospital admissions or medical records. Participants identified as having possible cognitive impairment undergo annual neuropsychological examinations until they either develop dementia, or two or more annual examinations suggest normality or show a reversion to previous cognitive functioning. Neurology examinations are also performed as indicated. An overall dementia diagnosis is based on whether criteria is met from the Diagnostic and Statistical Manual of Mental Disorders (IV-TR). For dementia cases that occurred prior to 2001, a re-review was conducted so that the latest diagnostic criteria was applied.

### Data Analyses

#### Association of AQP4 with Cognition and Brain Volumes

Statistical analyses were completed using the Statistical Analysis System (SAS) software v9.4. Only complete cases were included for analysis. Linear regression models were conducted to examine the relationship between *AQP4* (homozygote major [reference] versus heterozygote or versus homozygote minor) with cognition and brain volumes. To increase precision of estimates and reduce noise in the outcomes, all analyses were adjusted for age, age squared, sex, cohort (Original, Offspring, New Offspring Spouse, or Generation 3), *APOE* (ε4 carrier versus noncarrier), education (for cognitive outcomes; no high school degree, high school degree, some college, college graduate), and intracranial volume (for free water). Age squared was included because age has a non-linear association with cognition and brain volume. White matter hyperintensities volume, free water, and Trail Making Test B-A scores were natural log transformed because the data was not normally distributed. Higher scores were coded to reflect better cognitive performance across all measures (e.g., Trail Making Test scores were multiplied by negative 1 so higher scores indicated better performance).

#### Association of AQP4 with Dementia Risk

A Cox proportional hazard regression model was fitted to examine the association between *AQP4* with incident all-cause dementia risk. Cases were followed from baseline until the time they were diagnosed with dementia. Non-cases were censored at death or the last time point known to be dementia free, until 2022. The analysis was adjusted for age, sex, cohort (Original and Offspring), and *APOE* (ε4 carrier versus non-carrier). The proportionality of hazards assumption was met.

#### Replication in the UK Biobank

We sought to replicate our findings in an independent cohort by examining the association between *AQP4* with brain volumes and incident all-cause dementia risk in the UK Biobank. We did not include any cognitive outcomes because, despite the UK Biobank having cognitive data, the tests did not align with FHS cognitive domains or assessment modality (i.e., they were administered online). The UK Biobank is a prospective cohort study comprising 500,000 adults from the UK aged between 40 and 69 years at the time of recruitment from 2006 to 2010. Participants continue to be followed up for various health outcomes. See eMethods for further information regarding participants, MRI, and dementia case-ascertainment methods.

For *AQP4,* we only included rs335929 to represent *AQP4* because it was directly genotyped and had high linkage disequilibrium with all other *AQP4* haplotype SNPs (see eMethods).

Similar to FHS, linear regression models were conducted to examine the relationship between *AQP4* and brain volumes. The same brain volumes analyzed for FHS were included. Analyses were adjusted for age, age squared, sex, *APOE* (ε4 carrier versus non-carrier), ethnicity (white, other), MRI assessment centre (Cheadle, Reading, Newcastle, or Bristol), UK Biobank specific brain MRI adjustments (mean head motion, lateral brain position, transverse brain position, longitudinal brain position, and table position), and head scale (for free water). Brain volumes were multiplied by a head size scaling factor to standardize for individual differences in head size. White matter hyperintensities volume scores were natural log transformed because the data was not normally distributed. All brain MRI outcomes were converted to z-scores to enable direct comparison with the FHS results.

A Cox proportional hazard regression model was fitted to examine the association between *AQP4* with incident all-cause dementia risk to a subset of the UK Biobank cohort aged greater than 62.5 years at study entry to match the youngest participants included in the FHS analysis using all available follow-up. The analysis was adjusted for age, sex, *APOE* (ε4 carrier versus non-carrier), and ethnicity (white, other). The proportionality of hazards assumption was met. Analyses were performed using R software, version 4.2.3.

#### Pooled Analysis of AQP4 and Dementia Risk

An additional analysis was completed between *AQP4* and incident all-cause dementia risk using the pooled effects of FHS and UK Biobank samples using an inverse variance meta-analysis. Whilst results were considered significant if *p*<0.05, we did not account for multiple comparisons; instead, we primarily interpreted effect sizes and patterns of results.

#### Standard Protocol Approvals, Registrations, and Patient Consents

For the FHS, all participants provided written informed consent and ethical approval was obtained from the IRB at Boston University School of Medicine and the Human Research Ethics Committee at Monash University. For the UK Biobank, participants provided written informed consent and ethics approval was obtained from the North West Multi-Centre Research Ethics Committee as a Research Tissue Bank (RTB) approval. This means that the current study operates under RTB approval and a separate ethical clearance was not required.

#### Data Availability

The FHS makes phenotypic and genetic data available through online repositories BioLINCC and dbGAP, respectively. In addition, investigators can request data for specific projects through the FHS website for free: https://www.framinghamheartstudy.org/fhs-for-researchers/. UK Biobank data is available for a fee pending application approval from: https://www.ukbiobank.ac.uk/.

## RESULTS

### Characteristics of the FHS Samples

The characteristics of the cognition sample (N=3,847) are presented in Table 2. 65% of participants were homozygote major, 31% were heterozygote, and 4% were homozygote minor, which is broadly consistent with previous reports.^13^ The mean age of participants was 60 years; 53% were women. Characteristics of the MRI (N=3,332) and dementia (N=3,207) analysis samples are presented in eTables 1 and 2, respectively.

**Table 2.** Characteristics of the Cognition Sample in the FHS.

| AQP4 Haplotype Group | Homozygote Major | Heterozygote | Homozygote Minor | Overall |
| --- | --- | --- | --- | --- |
| N | 2513 | 1201 | 133 | 3847 |
| Age, years | 60.5 (10.8) | 60.9 (11.0) | 61.4 (11.2) | 60.6 (10.9) |
| Sex, women n (%) | 1342 (53.4) | 638 (53.1) | 76 (57.1) | 2056 (53.4) |
| Level of Education, n (%) |  |  |  |  |
| No high school | 81 (3.2) | 46 (3.8) | 5 (3.8) | 132 (3.4) |
| High school degree | 609 (24.2) | 294 (24.5) | 41 (30.8) | 944 (24.5) |
| Some college | 748 (29.8) | 380 (31.6) | 44 (33.1) | 1172 (30.5) |
| College degree | 1075 (42.8) | 481 (40.1) | 43 (32.3) | 1599 (41.6) |
| Clinical and Medical Information |  |  |  |  |
| Blood pressure (systolic), mmHg | 124 (18) | 125 (18) | 124 (16) | 124 (18) |
| Blood pressure (diastolic), mmHg | 74 (10) | 74 (10) | 73 (10) | 74 (10) |
| Prevalent Hypertension (stage 1), n (%) | 997 (39.8) | 504 (42.1) | 53 (39.9) | 1554 (40.5) |
| Prevalent Type 2 Diabetes, n (%) | 244 (10.0) | 115 (10.0) | 14 (10.7) | 373 (10.0) |
| Body Mass Index | 28.2 (5.6) | 28.0 (5.4) | 27.6 (4.5) | 28.129 (5.5) |
| Prevalent Cardiovascular disease, n (%) | 271 (10.8) | 139 (11.6) | 21 (15.8) | 431 (11.2) |
| Prevalent smoker, n (%) | 270 (10.8) | 124 (10.3) | 14 (10.5) | 408 (10.6) |
| Framingham Stroke Risk Profile (FSRP), score units | 0.042 (0.073) | 0.044 (0.072) | 0.045 (0.066) | 0.043 (0.072) |
| APOE ε4 carrier, n (%) | 562 (22.4) | 253 (21.1) | 28 (21.1) | 843 (21.9) |
| Cognitive Measures, n correct* |  |  |  |  |
| Logical Memory immediate recall | 11.7 (3.5) | 11.9 (3.4) | 11.6 (3.4) | 11.8 (3.5) |
| Logical Memory delayed recall | 10.7 (3.7) | 10.9 (3.7) | 10.8 (3.6) | 10.8 (3.7) |
| Visual Reproduction immediate recall | 8.8 (3.0) | 8.8 (3.1) | 8.4 (3.2) | 8.8 (3.0) |
| Visual Reproduction delayed recall | 8.0 (3.2) | 8.0 (3.3) | 7.5 (3.3) | 8.0 (3.3) |
| Trail Making Test B-A, completion time (seconds) | 0.9 (1.1) | 0.9 (1.1) | 0.9 (1.1) | 0.9 (1.1) |
| Similarities | 16.8 (3.5) | 16.6 (3.6) | 16.5 (3.5) | 16.7 (3.5) |

Note. Data are mean (SD) unless specified otherwise. \*except Trail Making Test B-A, which is measured in seconds.

### Association of AQP4 with Cognition and Brain Volumes

Compared to homozygote majors, heterozygotes displayed statistically significant higher Logical Memory immediate and delayed scores and larger hippocampal volumes (Table 3 and Table 4). To contextualize these effect sizes, in the same model, *APOE* ε4 carriers had statistically significant smaller hippocampal volumes compared to non-carriers (β[95% CI], -0.09 [-0.17, -0.01], *p*=0.029). Thus, the effect of *AQP4* on hippocampal volume was comparable to the magnitude of *APOE*, with an effect size ratio equal to 1.00. Associations between *AQP4* and the other brain MRI and cognitive outcomes were not significant.

**Table 3.** *AQP4* and Cognition in the FHS

|  | **N** | **β (95% CI )** | **P** |
| --- | --- | --- | --- |
| Logical Memory Immediate, n correct | 3847 |  |  |
| Homozygote Major | 2513 | REF |  |
| Heterozygote | 1201 | 0.26 (0.04, 0.48) | **0.018** |
| Homozygote Minor | 133 | 0.21 (-0.34, 0.77) | 0.453 |
| Logical Memory Delayed, n correct | 3847 |  |  |
| Homozygote Major | 2513 | REF |  |
| Heterozygote | 1201 | 0.32 (0.09, 0.55) | **0.007** |
| Homozygote Minor | 133 | 0.36 (-0.23, 0.95) | 0.229 |
| Visual Reproduction Delayed, n correct | 3847 |  |  |
| Homozygote Major | 2513 | REF |  |
| Heterozygote | 1201 | 0.04 (-0.15, 0.24) | 0.675 |
| Homozygote Minor | 133 | -0.28 (-0.78, 0.22) | 0.278 |
| Visual Reproduction Immediate, n correct | 3847 |  |  |
| Homozygote Major | 2513 | REF |  |
| Heterozygote | 1201 | -0.01 (-0.20, 0.17) | 0.893 |
| Homozygote Minor | 133 | -0.26 (-0.73, 0.21) | 0.277 |
| Trail Making Test B-A, time to completion* | 3838 |  |  |
| Homozygote Major | 2507 | REF |  |
| Heterozygote | 1198 | -0.001 (-0.02, 0.01) | 0.846 |
| Homozygote Minor | 133 | 0.02 (-0.02, 0.05) | 0.372 |
| Similarities, n correct | 3847 |  |  |
| Homozygote Major | 2513 | REF |  |
| Heterozygote | 1201 | -0.09 (-0.30, 0.13) | 0.427 |
| Homozygote Minor | 133 | 0.02 (-0.53, 0.56) | 0.957 |

*Note.* All analyses were adjusted for entry age, age squared, sex, education (no high school degree, high school degree, some college, college graduate), cohort (Original, Offspring, New Offspring, or Generation 3), and *APOE* (ε4 carrier versus non-carrier); Bold indicates statistical significance, $p < .05$ ; CI=confidence interval; \*reverse scored such that higher values indicate better performance. Values were also natural log transformed.

**Table 4.**
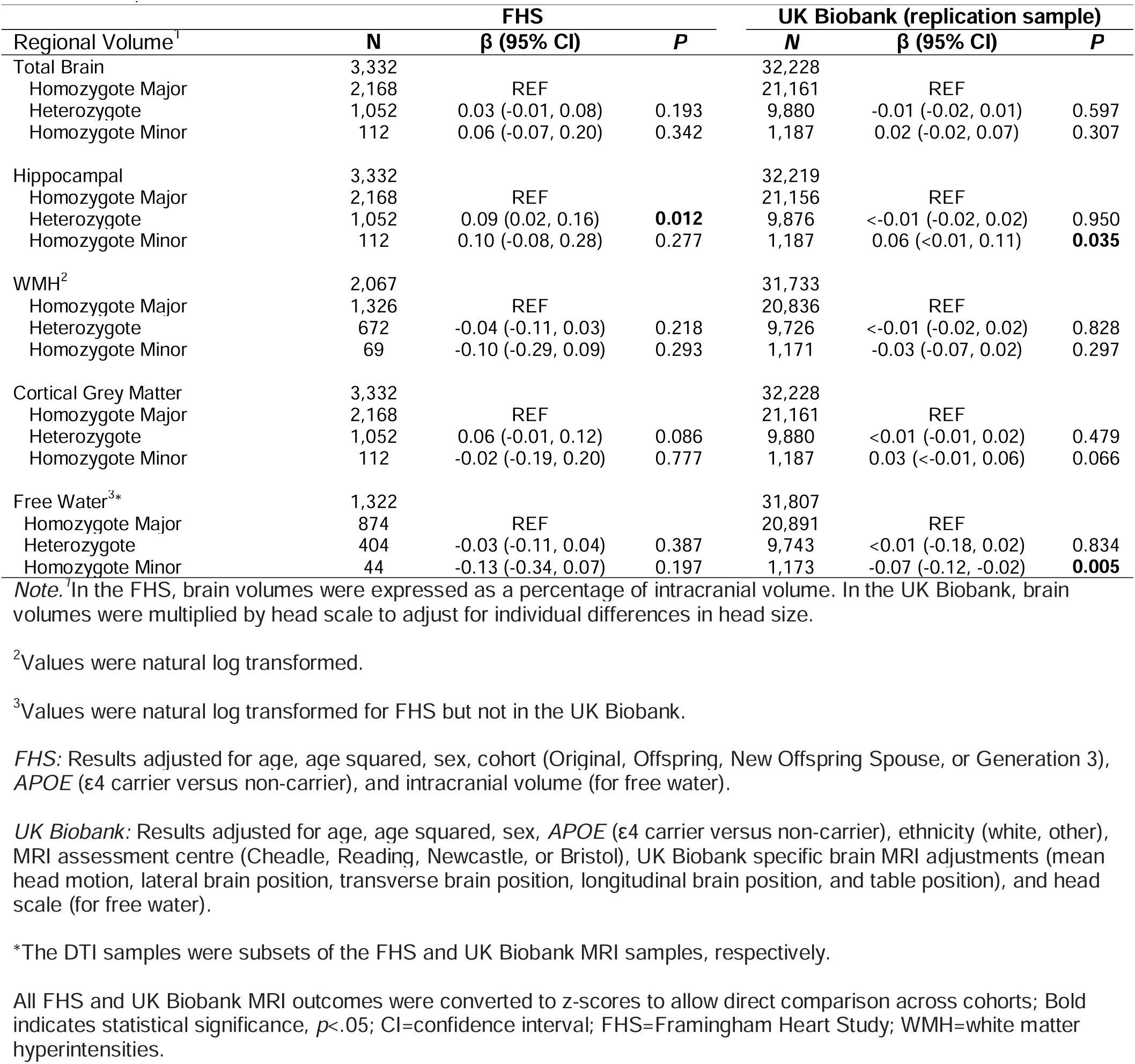
*AQP4* and Brain Volumes

### Association of AQP4 with Dementia Risk

The associations between *AQP4* and incident dementia risk are presented in Figure 1. Over an average of 20 years of follow-up (SD=7, maximum years of follow-up=40), there were 638 (20.0%) cases of incident dementia, 510 were due to clinical AD (80% of cases). Heterozygotes displayed a 12% lower rate of incident all-cause dementia compared to homozygote majors, although this did not reach significance. The association observed for homozygote minors, relative to homozygote majors, was nonsignificant and accompanied by wide confidence intervals, suggesting uncertainty regarding the size and direction of the associations. It should be noted that there were few events captured in the homozygote minor group (n events=31).

### Replication in the UK Biobank AQP4 and Brain Volumes

Characteristics of the UK Biobank brain MRI sample (N=32,228) are presented in eTable 3. Compared to homozygote majors, homozygote minors displayed statistically significant larger hippocampal volumes (Table 4). The magnitude of effect of *AQP4* on hippocampal volume was comparable to that of *APOE (*β[95% CI] = - 0.06[-0.08, -0.04], *p*=<0.01), with an effect size ratio equal to 1.00. The association between heterozygotes and hippocampal volume, relative to homozygote majors, was non-significant and accompanied by a small effect size. Homozygote minors displayed a statistically significant lower amount of brain free water compared to homozygote majors. Associations between *AQP4* and the other brain volumes were not significant.

### AQP4 and Dementia Risk

Characteristics of the UK Biobank dementia sample (N=114,868, incident cases=5,625) are presented in eTable 4. Compared to homozygote majors, heterozygotes displayed a statistically significant 7% lower rate of incident all-cause dementia over an average of 13 years of follow-up (SD=3, maximum years of follow-up=16; Figure 1). The association between homozygote minors and dementia risk, relative to homozygote majors, was nonsignificant and close to null.

### Pooled Dementia Analysis

Across cohorts (N=118,075, incident cases=6,263), heterozygotes displayed a statistically significant 7% lower rate of incident all-cause dementia (Figure 1). The association between homozygote minors and dementia risk, relative to homozygote majors, was nonsignificant and close to the null.

## DISCUSSION

We found that, relative to homozygote majors, heterozygotes displayed better verbal episodic memory and larger hippocampal volumes in the FHS. In the UK Biobank, homozygote minors had larger hippocampal volumes and a lower amount of brain free water compared to homozygote majors. Across both samples, heterozygotes displayed a 7% lower rate of incident dementia. Overall, these results suggest a beneficial effect of carrying one minor allele.

Few studies have investigated the association between *AQP4* and brain health. In relation to cognition, one study revealed a potential protective effect of carrying at least one minor allele at the *AQP4* haplotype, demonstrated by better reaction times following sleep deprivation in healthy minor allele carriers versus non-carriers.^17^ Another found carrying the minor allele at rs335929, one of the *AQP4* haplotype SNPs, was associated with slower decline in Logical Memory and Digit Span performance, but more rapid functional decline in individuals with AD dementia.^15^ Our findings also support the notion that, in persons without dementia, the minor allele is associated with better memory.

To our knowledge, no previous studies have investigated the relationship between *AQP4* and brain volume in humans. However, studies conducted on mice suggest *AQP4* plays a role in learning and memory,^33,34^ which may be subserved by the hippocampus. Specifically, *AQP4* knockout mice display reduced long-term potentiation and synaptic plasticity in the hippocampi.^18,35,36^ Additionally, knockout mice have reduced performance on a spatial memory task compared to wild type mice,^37^ highlighting the potential role AQP4 has in hippocampal-dependent learning and memory. In our study, the effect of *AQP4* on hippocampal volume was comparable to the effect of *APOE* in both the FHS and UK Biobank samples.

We also found some evidence to suggest that homozygote minors had lower amounts of brain free water compared to homozygote majors. Interestingly, a previous study also found an association between *AQP4* SNPs not included in our study and free water.^38^ Differences in free water by *AQP4* align with the role of AQP4 in fluid transport and may reflect homozygote minors having less extracellular fluid accumulation or healthier myelin. However, further studies are needed to explore this.

We found a 7% lower rate of incident dementia for heterozygotes compared to homozygote majors, which is broadly consistent with the outcomes discussed above. A previous genome-wide association study (GWAS) identified that a SNP in the adjacent *AQP4* antisense gene (*AQP4-AS1*; rs187423924) was associated with late-onset Alzheimer’s disease.^39^ Although this variant is not in strong linkage disequilibrium with the SNPs evaluated in our haplotype, the presence of independent associations within the *AQP4* genomic region supports the broader hypothesis that AQP4 pathways may contribute to dementia risk. Our findings add support to this hypothesis by demonstrating that variation in an *AQP4* haplotype is associated with cognition, MRI outcomes, and incident dementia.

A number of underlying mechanisms may explain why carrying the minor allele at this *AQP4* haplotype is associated with better brain-related outcomes. Firstly, carrying the minor allele at the haplotype has been associated with reduced AQP4 channel expression compared to homozygote majors.^18^ Interestingly, Larsen et al^17^ found carrying the *AQP4* minor allele was associated with increased slow-wave energy (a measure of the power of slow wave oscillations in the EEG waveform) during NREM sleep, particularly early in the night. It was proposed that higher levels of slow wave energy may compensate for having a reduced expression of AQP4 channels, considering glymphatic clearance, including clearance of Aβ and tau, is thought to be highly coupled to slow wave sleep.^40–42^ Increased slow wave energy facilitates learning and memory consolidation ^43,44^ as well as memory performance in the face of high Aβ burden.^45^ Variation in AQP4 channel expression may also have implications for BBB permeability and integrity. For instance, it has been suggested that AQP4 expression can be up or down regulated following BBB disruption^46–48^ and perhaps *AQP4* genetic variation influences the brain’s response to BBB injury and neuroinflammation. Further research is needed to explore additional mechanisms that may impact these relationships. For example, investigating whether *AQP4* influences Aβ and tau dynamics and the potential moderation age, biological sex, and sleep may have on this relationship.

We did not observe a general dose-response relationship for *AQP4* across the outcomes examined. It is possible that the lower proportion of participants in the homozygote minor group may have limited the power to detect an effect in this group, particularly in the FHS sample. For example, when considering the pooled effect for incident dementia, the confidence intervals for homozygote minors encapsulated the point estimate for heterozygotes. It is therefore possible that *AQP4* may have a non-linear relationship, in which carrying one or two minor alleles produces comparable effects. Other non-additive genetic effects are not uncommon. For example, heterozygosity for *Klotho* (KL) has been associated with better health outcomes, including lower dementia risk, as compared to being homozygote for the major or minor allele.^49^ It is also possible that the associations for each genotype may vary by outcome, whereby each outcome has its own distinct biological pathway. Additional studies will be required to explore these ideas further.

## Strengths and Limitations

Strengths of our study include the large sample of community-dwelling adults followed prospectively and replication in a large independent sample. Both cohorts primarily included individuals of European backgrounds, limiting the generalizability of the study’s findings to other racial or ethnic groups. Although this study provides an important first step, future studies will be required to replicate these results in more diverse samples and explore whether *AQP4* genetic variation relates to different neuropathological substrates.

## Conclusion

We found that carrying the minor allele at an *AQP4* haplotype was associated with better verbal episodic memory, larger hippocampal volumes, lower amounts of brain free water, and lower dementia risk. These findings reveal a potential protective effect of the minor allele at this *AQP4* haplotype. Continuing to explore the potential role AQP4 has in dementia may be important in elucidating additional processes that could contribute to the onset and progression of AD and related dementias.

## Acknowledgements

We would like to thank the Framingham Heart Study and UK Biobank participants for their commitment and dedication to their respective studies. This research has been conducted using the UK Biobank Resource under application number 70607.

