## Supplementary Materials for "Association of an Aquaporin-4 Haplotype with Cognition, Brain Volume, and Dementia Risk in Community-Dwelling Individuals without Dementia"

### Contents

eMethods. UK Biobank Methods: Participants

eMethods. UK Biobank Methods: Brain MRI Protocol

eMethods. UK Biobank Methods: Dementia Case Ascertainment

eMethods. UK Biobank Methods: Genotyping of AQP4

eMethods. UK Biobank Methods: Genotyping of *AQP4*

eFigure 1. Cognitive Sample Selection in the FHS

eFigure 2. Brain MRI Sample Selection in the FHS

eFigure 3. Incident Dementia Sample Selection in the FHS

eTable 1. Characteristics of the Brain MRI Sample in the FHS

eTable 2. Characteristics of the Dementia Analysis Sample in the FHS

eTable 3. Characteristics of the Brain MRI Sample in the UK Biobank

eTable 4. Characteristics of the Dementia Sample in the UK Biobank

eFigure 4. Brain MRI Sample Selection in the UK Biobank

eFigure 5. Dementia Sample Selection in the UK Biobank

eReferences

### eMethods

#### UK Biobank Methods: Participants

Participants were excluded who had dementia or another significant neurological condition at baseline or the time of MRI. Sample selection information is presented in eFigures 4 and 5.

#### UK Biobank Methods: Brain MRI Protocol

From 2014 to 2023, approximately 100,000 participants completed the UK Biobank's imaging sub-study. Brain MRIs were completed on 3T Siemens Skra scanners. Further information regarding MRI procedures has been published previously.[1]

#### UK Biobank Methods: Dementia Case Ascertainment

Within the UK Biobank, information is available on incident all-cause dementia. Participant's dementia status was determined from hospital records, death records, and primary care providers.[2] The algorithms used to calculate all-cause dementia were created to maximise positive predictive value (>80% for each record type).[2] Dementia diagnoses were based on Read V2 (primary care) and ICD-10 codes.[2] Follow-up for dementia status in the current study was available up until 2022.

#### UK Biobank Methods: Genotyping of *AQP4*

All participants had genome-wide genotype data collected at baseline. Genotyping was completed at the ARSL in Santa Clara, California, USA and conducted on peripheral blood samples stored at -80°C using the UK Biobank Axiom Array. Using this array, approximately 850,000 variants were directly measured and>90 million variants were imputed using the Haplotype Reference Consortium and UK10K + 1000 Genomes reference panels. Allele calling was completed with Affymetrix Power Tools Algorithm. The following quality-control criteria were applied by the UK Biobank data management team: minor allele frequency (MAF)≥0.01, HWE of $p \geq 10^{-6}$, and call rate≥0.95. A principal-components analysis was applied to evaluate the population structure using a set of 407,219 unrelated samples, revealing that the majority of participants were British (88.26%).[3] Further information regarding genetic information in the UK Biobank has been published previously.[3]

For *AQP4*, we included rs335929 to represent the *AQP4* haplotype. It has high linkage disequilibrium with all other SNPs in the haplotype across different samples, including the UK Biobank as shown from: https://snipa.org/snipa3/index.php?task=pairwise_ld(LD r 2 >.87; Genome assembly=GRCh37; variant set: 1000 Genomes, Phase 3 v 5; population=European (78% of UK biobank participants have European ancestry); genome annotation=Ensembl 87; LD r 2 threshold=0.8).

**Linkage Disequilibrium of SNPs in the *AQP4* Haplotype**

| SNPs | LD $R^2$ |
|---|---|
| rs335929 and rs335930 | .87 |
| rs335929 and rs162007 | .91 |
| rs335929 and rs162008 | .91 |
| rs335929 and rs63514 | .95 |
| rs335929 and rs335931 | .97 |
| rs335929 and rs16942851 | .99 |
| rs335929 and rs455671 | 1 |

*Note.* SNPs=single nucleotide polymorphisms; LD=linkage disequilibrium.

**eFigure 1. Cognitive Sample Selection in the FHS**

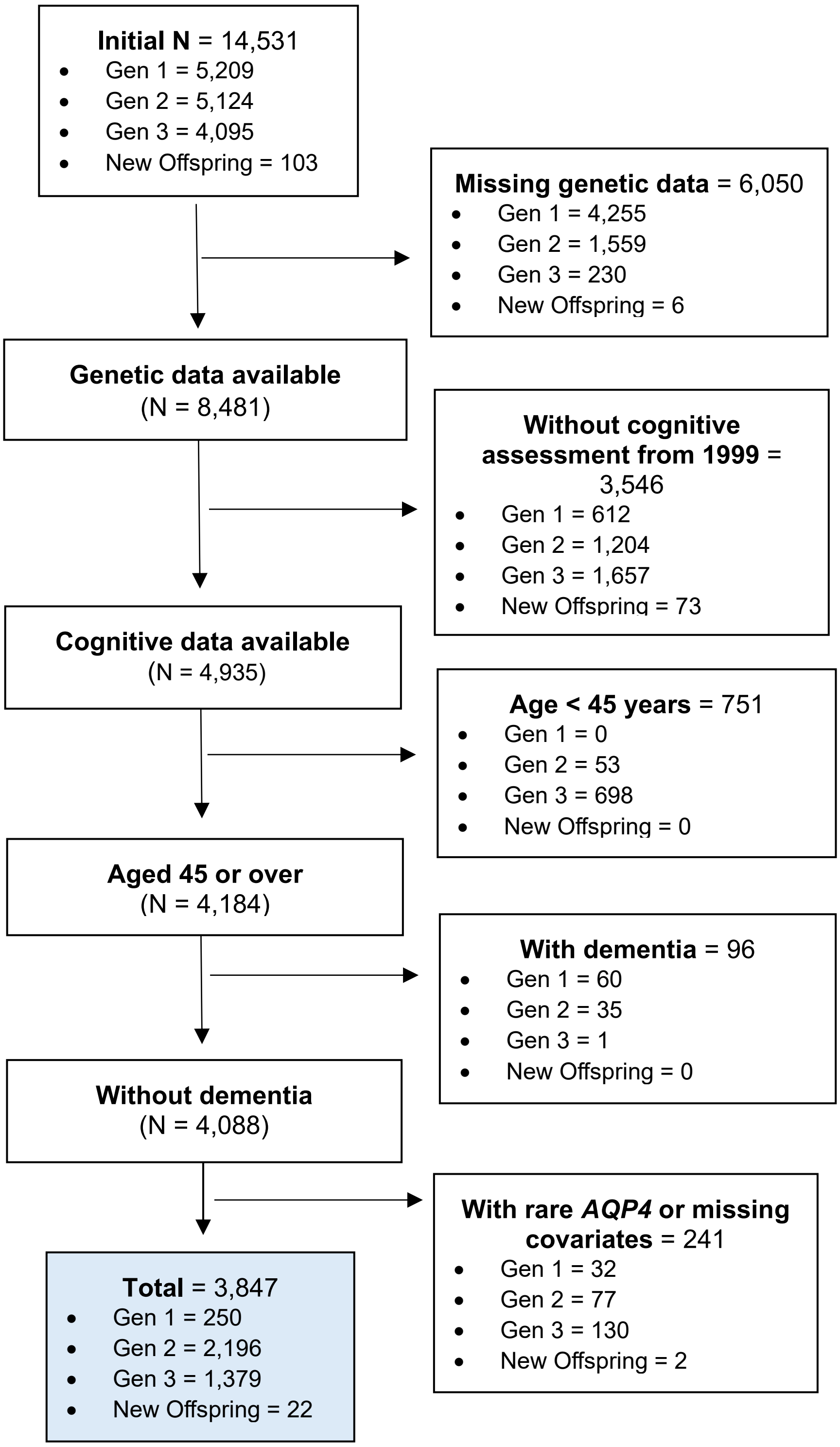

**eFigure 2. Brain MRI Sample Selection in the FHS.** DTI = Diffusion Tensor Imaging

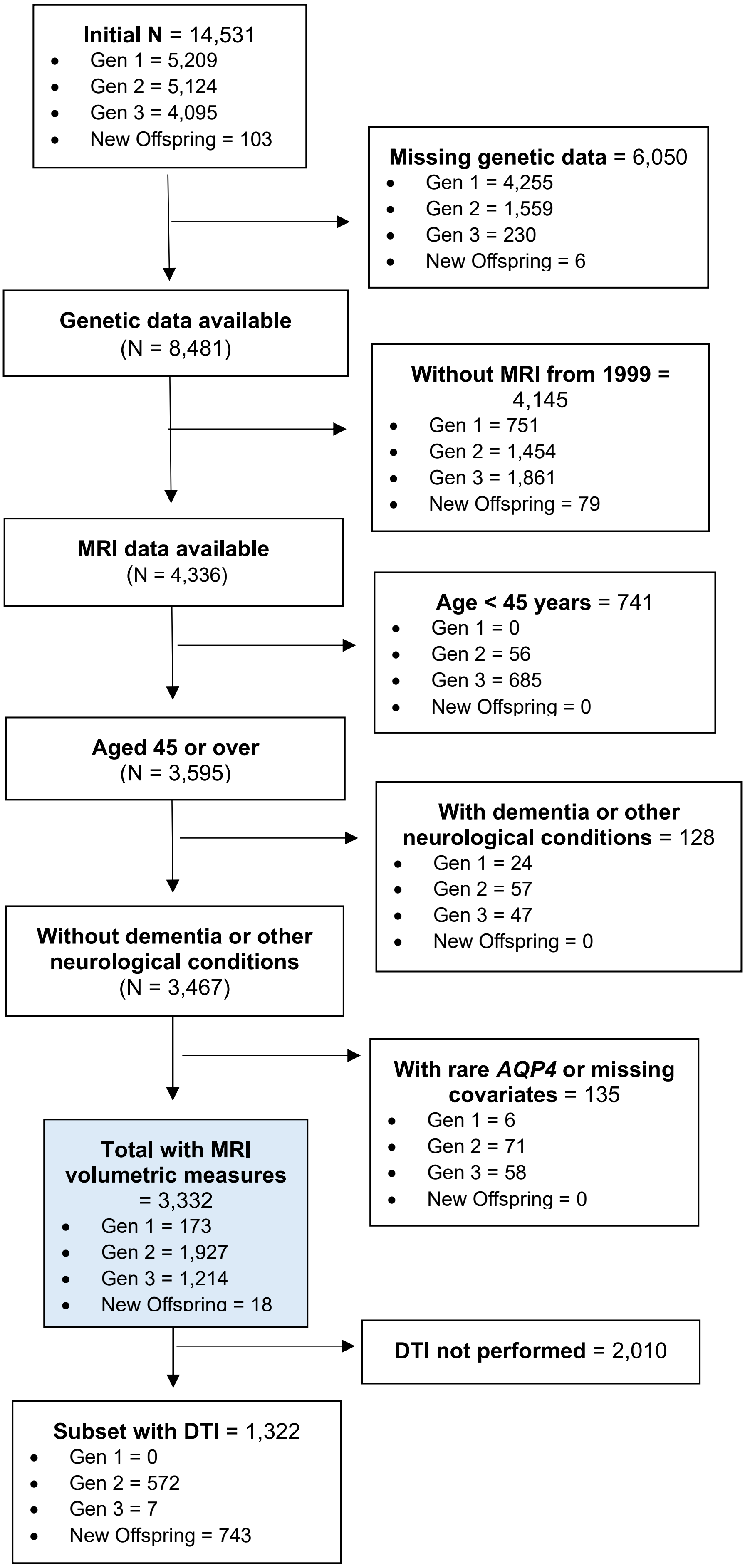

**eFigure 3. Incident Dementia Sample Selection in the FHS**

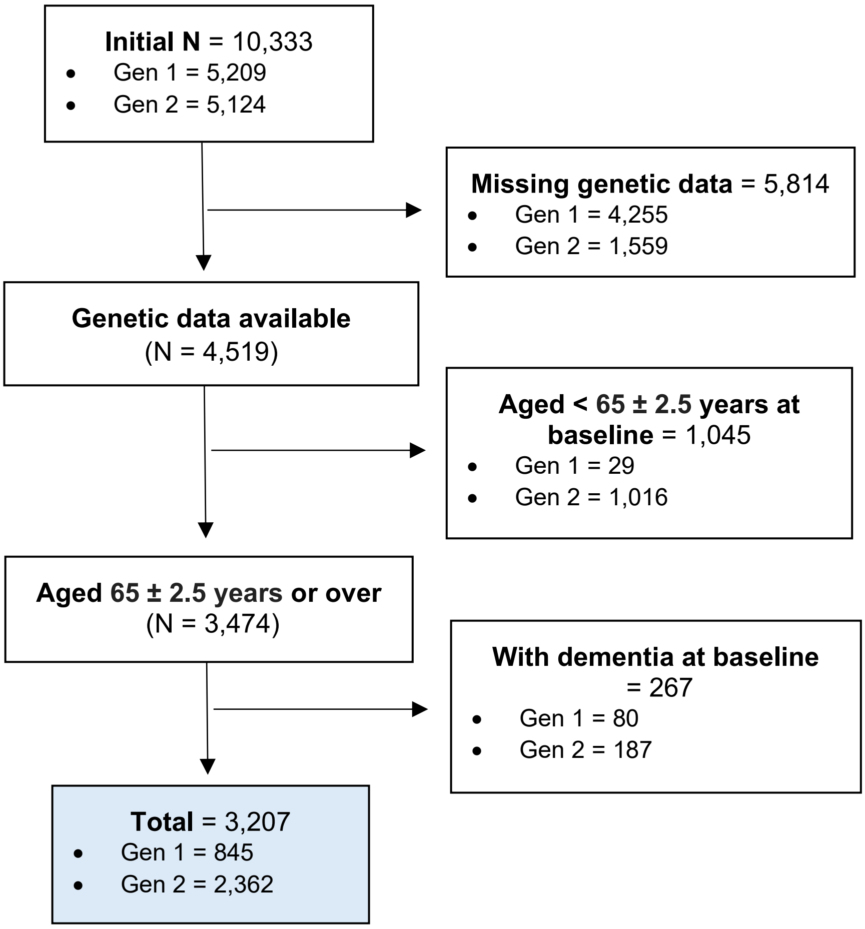

**eTable 1. Characteristics of the Brain MRI Sample in the FHS**

| *AQP4* Haplotype Group | Homozygote Major Allele Carriers | Heterozygote Carriers | Homozygote Minor Allele Carriers | Overall |
|---|---|---|---|---|
| N | 2168 | 1052 | 112 | 3332 |
| Age, years | 60.2 (10.4) | 60.6 (10.6) | 60.5 (10.8) | 60.4 (10.5) |
| Women, n (%) | 1154 (53.2) | 553 (52.6) | 60 (53.6) | 1767 (53.0) |
| Level of Education, n (%) | | | | |
| No high school | 64 (3.0) | 36 (3.4) | 5 (4.5) | 105 (3.2) |
| High school degree | 508 (23.4) | 245 (23.3) | 33 (29.5) | 786 (23.6) |
| Some college | 637 (29.4) | 343 (32.6) | 36 (32.1) | 1016 (30.5) |
| College degree | 959 (44.2) | 428 (40.7) | 38 (33.9) | 1425 (42.8) |
| Clinical and Medical Information | | | | |
| Blood pressure (systolic), mmHg | 124 (18) | 124 (18) | 125 (16) | 124 (18) |
| Blood pressure (diastolic), mmHg | 74 (10) | 74 (10) | 73 (10) | 74 (10) |
| Prevalent Hypertension (stage 1), n (%) | 832 (38.4) | 419 (40.0) | 43 (38.7) | 1294 (38.9) |
| Prevalent Type 2 Diabetes, n (%) | 218 (10.3) | 99 (9.7) | 11 (9.9) | 328 (10.1) |
| Body Mass Index | 28.1 (5.4) | 27.8 (5.3) | 27.5 (4.5) | 28.0 (5.3) |
| Prevalent Cardiovascular disease, n (%) | 210 (9.7) | 106 (10.1) | 12 (10.7) | 328 (9.8) |
| Prevalent smoker, n (%) | 229 (10.6) | 106 (10.1) | 13 (11.6) | 348 (10.5) |
| Framingham Stroke Risk Profile (FSRP), score units | 0.040 (0.069) | 0.042 (0.066) | 0.040 (0.057) | 0.041 (0.068) |
| *APOE* ε4 carrier, n (%) | 485 (22.37) | 224 (21.29) | 27 (24.11) | 736 (22.09) |
| MRI outcomes, $cm^3$ | | | | |
| Total Cerebral Brain Volume | 982.9 (105.6) | 982.0 (109.2) | 968.5 (100.3) | 982.2 (106.6) |
| Hippocampal Volume | 6.7 (0.7) | 6.7 (0.8) | 6.6 (0.7) | 6.7 (0.8) |
| White Matter Hyperintensities Volume | 1.7 (3.9) | 1.6 (2.9) | 1.8 (2.7) | 1.7 (3.6) |
| Cortical Grey Matter | 487.6 (50.0) | 487.6 (50.7) | 478.9 (45.8) | 487.3 (50.1) |
| Free Water, volume fraction | 0.21 (0.04) | 0.21 (0.03) | 0.22 (0.04) | 0.21 (0.04) |

*Note.* Data are mean (SD) unless specified otherwise; ICV=intracranial volume.

**eTable 2. Characteristics of the Dementia Sample in FHS**

| *AQP4* Haplotype Group | Homozygote Major Allele Carriers | Heterozygote Carriers | Homozygote Minor Allele Carriers | Overall |
|---|---|---|---|---|
| N | 2,094 | 988 | 125 | 3,207 |
| Age, years | 64.9 (1.1) | 64.9 (1.1) | 65.0 (1.1) | 64.9 (1.1) |
| Women, n (%) | 1,176 (56.2) | 554 (56.1) | 72 (57.6) | 1,802 (56.2) |
| Level of Education, n (%) | | | | |
| No high school | 258 (12.5) | 119 (12.2) | 11 (8.9) | 338 (12.2) |
| High school degree | 684 (33.0) | 327 (33.5) | 46 (37.1) | 1,057 (33.3) |
| Some college | 530 (25.6) | 255 (26.1) | 38 (30.6) | 823 (26.0) |
| College degree | 598 (28.9) | 276 (28.3) | 29 (23.4) | 903 (28.5) |
| Clinical and Medical Information | | | | |
| Blood pressure (systolic), mmHg | 132 (19) | 132 (18) | 131 (17) | 132 (18) |
| Blood pressure (diastolic), mmHg | 76 (10) | 77 (10) | 77 (10) | 76 (10) |
| Prevalent Hypertension (stage 1), n (%) | 1,115 (53.5) | 552 (56.0) | 65 (52.0) | 1,732 (54.2) |
| Prevalent Type 2 Diabetes, n (%) | 201 (10.4) | 112 (12.2) | 13 (11.5) | 326 (11.0) |
| Waist to Hip Ratio | 0.9 (0.1) | 0.9 (0.1) | 0.9 (0.1) | 0.9 (0.1) |
| Prevalent Cardiovascular disease, n (%) | 289 (13.8) | 134 (13.6) | 26 (20.8) | 449 (14.0) |
| Prevalent smoker, n (%) | 289 (14.1) | 135 (13.8) | 21 (16.9) | 445 (14.1) |
| Framingham Stroke Risk Profile (FSRP), score units | 0.042 (0.034) | 0.044 (0.036) | 0.043 (0.042) | 0.042 (0.035) |
| *APOE* ε4 carrier, n (%) | 470 (22.5) | 211 (21.4) | 26 (20.8) | 707 (22.1) |

*Note.* Data are mean (SD) unless specified otherwise.

**eTable 3. Characteristics of the Brain MRI Sample in the UK Biobank**

| *AQP4* Haplotype Group | Homozygote Major Allele Carriers | Heterozygote Carriers | Homozygote Minor Allele Carriers | Overall |
|---|---|---|---|---|
| N | 21,161 | 9,880 | 1,187 | 32,228 |
| Age, years | 64.0 (7.7) | 64.0 (7.8) | 63.8 (7.8) | 64.0 (7.7) |
| Women, n (%) | 11,301 (53.4) | 5,187 (52.5) | 656 (55.3) | 17,144 (53.2) |
| Level of Education, n (%) | | | | |
| A levels | 1,251 (5.9) | 615 (6.2) | 68 (5.7) | 1,934 (6.0) |
| CSE or O levels | 2,727 (12.9) | 1,278 (12.9) | 156 (13.1) | 4,161 (12.9) |
| NVQ or HND or HNC | 2,556 (12.1) | 1,129 (11.4) | 132 (11.1) | 3,817 (11.8) |
| Progressional qual or graduate | 13,122 (62.0) | 6,171 (62.5) | 766 (64.5) | 20,059 (62.2) |
| None of the above | 1,304 (6.2) | 590 (6.0) | 59 (5.0) | 1,953 (6.1) |
| Prefer not to answer | 68 (0.3) | 28 (0.3) | 1 (0.1) | 97 (0.3) |
| Ethnicity, n (%) | | | | |
| White | 19,585 (92.6) | 9,076 (91.9) | 1,080 (91.0) | 29,741 (92.3) |
| Other | 1,506 (7.1) | 781 (7.9) | 107 (9.0) | 2,394 (7.4) |
| Clinical and Medical Information | | | | |
| Blood pressure (systolic), mmHg | 139 (19) | 139 (18) | 138 (19) | 139 (19) |
| Blood pressure (diastolic), mmHg | 79 (10) | 79 (10) | 78 (10) | 79 (10) |
| Prevalent Hypertension, n (%) | 7,879 (37.2) | 3,583 (36.3) | 389 (32.8) | 11,851 (36.8) |
| Prevalent Type 2 Diabetes, n (%) | 1,032 (4.9) | 469 (4.8) | 49 (4.1) | 1,550 (4.8) |
| Body Mass Index | 26.47 (4.3) | 26.41 (4.3) | 26.40 (4.4) | 26.45 (4.3) |
| Prevalent Cardiovascular disease, n (%) | 1,432 (6.8) | 682 (6.9) | 75 (6.3) | 2,189 (6.8) |
| Prevalent smoker, n (%) | 635 (3.0) | 331 (3.4) | 45 (3.8) | 1,011 (3.1) |
| *APOE* ε4 carrier, n (%) | 5,903 (27.9) | 2,701 (27.3) | 340 (28.6) | 8,944 (27.8) |
| MRI outcomes, $cm^3$ | | | | |
| Total Cerebral Brain Volume | 1490.6 (73.7) | 1490.6 (74.1) | 1493.4 (74.8) | 1490.7 (73.9) |
| Hippocampal Volume Total | 9.9 (1.2) | 9.9 (1.2) | 10.0 (1.2) | 9.9 (1.2) |
| White Matter Hyperintensities Volume | 5.1 (6.6) | 5.1 (6.7) | 4.8 (6.1) | 5.1 (6.7) |
| Cortical Grey Matter Volume | 545.1 (61.8) | 544.1 (62.1) | 541.3 (61.0) | 544.7 (61.8) |
| Free Water, volume fraction | 0.09 (0.01) | 0.09 (0.01) | 0.09 (0.01) | 0.09 (0.01) |
| Adjustments | | | | |
| Head motion | 0.2 (0.1) | 0.2 (0.1) | 0.2 (0.1) | 0.2 (0.1) |
| Lateral brain position | 0.8 (3.0) | 0.8 (3.1) | 0.8 (3.2) | 0.8 (3.1) |
| Transverse brain position | 66.5 (5.9) | 66.4 (5.9) | 66.5 (6.2) | 66.5 (5.9) |
| Longitudinal brain position | -28.4 (26.5) | -28.7 (26.6) | -28.2 (26.3) | -28.5 (26.5) |
| Table position | -1054.1 (24.9) | -1054.0 (24.9) | -1054.0 (24.4) | -1054.0 (24.9) |
| Head scale | 1.3 (0.1) | 1.3 (0.1) | 1.3 (0.1) | 1.3 (0.1) |
| Assessment center, n (%) | | | | |
| Cheadle | 12,405 (58.6) | 5,883 (59.5) | 701 (59.1) | 18,989 (58.9) |
| Reading | 3,200 (15.1) | 1,538 (15.6) | 195 (16.4) | 4,933 (15.3) |
| Newcastle | 5,526 (26.1) | 2,448 (24.8) | 290 (24.4) | 8,264 (25.6) |
| Bristol | 30 (0.1) | 11 (0.1) | 1 (0.1) | 42 (0.1) |

*Note.* Data are mean (SD) unless specified otherwise; A levels=Advanced Level qualifications; CSE or O levels=Certificate of Secondary Education or Ordinary Level; NVQ or HND or HNC=National Vocational Qualification, Highter National Diploma, or Higher National Certificate; Professional qual or graduate=professional qualifications such as a certificate or a higher educational degree.

**eTable 4. Characteristics of the Dementia Sample in the UK Biobank**

| *AQP4* haplotype Group | Homozygote Major Allele Carriers | Heterozygote Carriers | Homozygote Minor Allele Carriers | Overall |
|---|---|---|---|---|
| N | 75,714 | 34,936 | 4,218 | 114,868 |
| Age, years | 65.8 (2.0) | 65.8 (2.0) | 65.9 (2.0) | 65.8 (2.0) |
| Women, n (%) | 39,548 (52.2) | 18,368 (52.6) | 2,228 (52.8) | 60,144 (52.4) |
| Level of Education, n (%) | | | | |
| A levels | 3,291 (4.4) | 1,507 (4.3) | 224 (5.3) | 5,022 (3.5) |
| CSE or O levels | 11,865 (15.7) | 5,407 (15.5) | 659 (15.6) | 17,931 (15.6) |
| NVQ or HND or HNC | 7,024 (9.3) | 3,226 (9.2) | 346 (8.2) | 10,596 (9.2) |
| Progressional qual or graduate | 30,103 (39.8) | 14,020 (40.1) | 1,751 (41.5) | 45,874 (39.9) |
| None of the above | 22,261 (29.4) | 10,224 (29.3) | 1,161 (27.5) | 33,646 (29.3) |
| Prefer not to answer | 1,121 (1.5) | 524 (1.5) | 75 (1.8) | 1,720 (1.5) |
| Ethnicity, n(%) | | | | |
| White | 70,724 (93.4) | 32,530 (93.1) | 3,861 (91.5) | 107,115 (93.3) |
| Other | 4,990 (6.6) | 2,406 (6.9) | 357 (8.5) | 7,753 (6.8) |
| Clinical and Medical Information | | | | |
| Blood pressure (systolic), mmHg | 145 (19) | 145 (19) | 145 (18) | 145 (19) |
| Blood pressure (diastolic), mmHg | 82 (10) | 82 (10) | 82 (10) | 82 (10) |
| Prevalent Hypertension, n (%) | 28,936 (38.2) | 13,330 (38.2) | 1,644 (39.0) | 43,910 (38.2) |
| Prevalent Type 2 Diabetes, n (%) | 5,660 (7.5) | 2,466 (7.1) | 336 (8.0) | 8,462 (7.4) |
| Body Mass Index | 27.55 (4.4) | 27.47 (4.4) | 27.52 (4.5) | 27.52 (4.4) |
| Prevalent Cardiovascular disease, n (%) | 32,603 (43.1) | 15,016 (43.0) | 1,832 (43.4) | 49,451 (43.1) |
| Prevalent smoker, n (%) | 5,327 (7.0) | 2,602 (7.5) | 292 (6.9) | 8,221 (7.2) |
| *APOE* ε4 carrier, n (%) | 21,486 (28.4) | 9,855 (28.2) | 1,179 (28.0) | 32,520 (28.3) |

*Note.* Data are mean (SD) unless specified otherwise; A levels=Advanced Level qualifications; CSE or O levels=Certificate of Secondary Education or Ordinary Level; NVQ or HND or HNC=National Vocational Qualification, Highter National Diploma, or Higher National Certificate; Professional qual or graduate=professional qualifications such as a certificate or a higher educational degree.

**eFigure 4. Brain MRI Sample Selection in the UK Biobank.** DTI = Diffusion Tensor Imaging

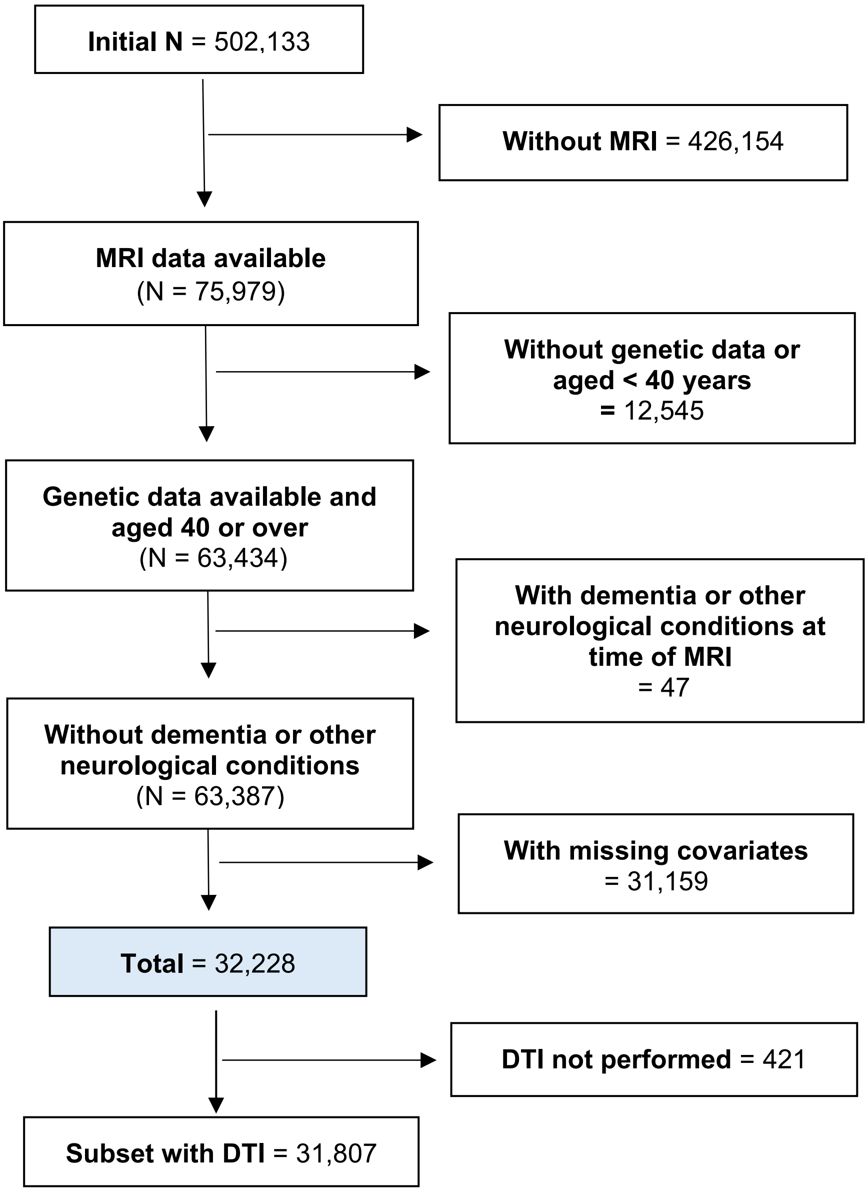

**eFigure 5. Dementia Sample Selection in the UK Biobank**

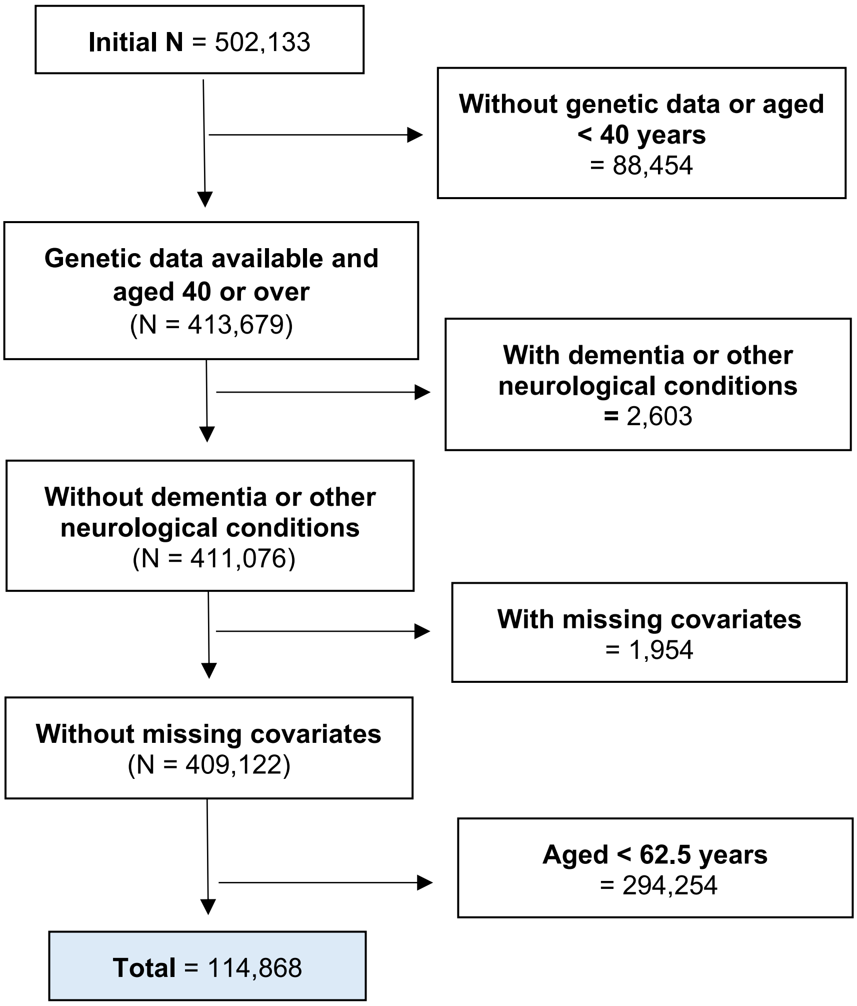
